# Genetic association of primary non-response to Anti-TNFα therapy in patients with Inflammatory Bowel Disease

**DOI:** 10.1101/2020.06.23.20138495

**Authors:** T. De, H. Zhang, C. Alarcon, B. Lec, J. Avitia, E. Smithberger, Chuyu Chen, M. Horvath, S. Kwan, M. Young, S. Adhikari, J. H. Kwon, J. Pacheco, G.P. Jarvik, W. Wei, F. Mentch, H. Hakonarson, P. Sleiman, A.S. Gordon, J. Harley, J. G. Linneman, S. Hebbring, L. Parisiadou, M. A. Perera

## Abstract

**Background & Aims:** Primary non-response (PNR) to anti-tumor necrosis factor-α (TNFα) biologics is a serious concern in patients with inflammatory bowel disease (IBD). We aimed to identify the genetic variants associated with PNR.

**Methods:** Patients were recruited from outpatient GI clinics and PNR was determined using both clinical and endoscopic findings. A case-control genome-wide association study was performed in 589 IBD patients and associations were replicated in an independent cohort of 293 patients. Effect of the associated variant on gene expression and TNFα secretion was assessed by cell-based assays. Pleiotropic effects were investigated by Phenome-wide Association Study (PheWAS).

**Results:** We identified rs34767465 as associated with PNR to anti-TNF-α therapy (OR:2.07, 95%CI:1.46-2.94, p=2.43×10^−7^, [Replication OR:1.8, 95%CI:1.04-3.16, p=0.03]). rs34767465 is a multiple-tissue expression quantitative trait loci for *FAM114A2*. Using RNA-sequencing and protein quantification from HapMap lymphoblastoid cell lines (LCLs), we found a significant decrease in *FAM114A2* mRNA and protein expression in both heterozygous and homozygous genotypes when compared to wild type LCLs. TNF-α secretion was significantly higher in THP-1 cells [differentiated into macrophages] with *FAM114A2* knockdown versus controls. Immunoblotting experiments showed that depletion of *FAM114A2* impaired autophagy related pathway genes suggesting autophagy mediated TNF-α secretion as a potential mechanism. PheWAS showed rs34767465 was associated with comorbid conditions found in IBD patients (derangement of joints [p=2.3×10^−4^], pigmentary iris degeneration [p=5.1×10^−4^], diverticulum of esophagus [p=6.3×10^−4^]).

**Conclusion:** We identified a variant rs34767465 associated with PNR to anti-TNFα biologics, which increases TNFα secretion through mechanism related to autophagy. rs34767465 may also explain the comorbidities associated with IBD.

## Introduction

Anti-tumor necrosis factor-alpha (anti-TNFα) biologics have transformed the therapeutic landscape of inflammatory bowel disease (IBD) in achieving clinical remission and endoscopic healing, and decreasing the risk of surgery, hospitalization and disease-related complications.^1^ In acute settings approximately 60-75% of patients who receive anti-TNF therapy achieve a clinical response and 20-40% of patients achieve a clinical remission.^1^ However, 30-40% of patients may not respond [primary non-responders] to these medications.^2^ Primary non-response (PNR) to anti-TNFα biologics has serious implications on disease course, including reduced likelihood of response to a second anti-TNF agent, and a greater need for surgery.^3^ Moreover, these agents are expensive and are associated with significant risks of infections and autoimmune complications.^4^ Therefore, identifying the factors that predict efficacy is crucial to allow clinicians to effectively use these biologics.

To date, limited studies have focused on identifying the risk factors associated with PNR to anti-TNFα therapy. Clinical risk factors like age, disease-associated features, body mass index, disease severity and extent associated with PNR^5^ have unclear predictive ability and remain poorly replicated, thereby limiting utility in clinical practice. Several studies have tested the association between C-reactive protein (CRP) level, a biomarker of inflammation and response to anti-TNF. While Arnott *et*.*al*., observed no association between CRP level and response,^6^ other studies observed a decrease of 25% or more in CRP level was associated with beneficial response.^7–9^ TNFα level has also been investigated as biomarker for anti-TNF therapy. In a small study of 36 patients with Crohn’s disease (CD), PNR was associated with higher baseline TNFα levels.^10^ However, findings could not be replicated in larger studies.^11, 12^ Similarly, Anti-Neutrophil Cytoplasmic Antibodies (ANCA), a family of antibodies related to inflammatory diseases have been the subject of several investigations for its association with anti-TNF response and have conflicting findings.^11^

Few studies investigated the genetic profiles of primary non-responders by targeting genes related to cytokines and their receptors or immunoglobulin receptors. The association of individual candidate variants within IL-13 receptor (IL13Rα2), IL-23 receptor (IL23R), TNF-receptor I (TNFRI), neonatal Fc receptor (VNTR2/VNTR3), apoptosis-related genes (Fas ligands, caspase 9), IgG Fc receptor IIIa (FcYRIIIa), and MAP kinases have been evaluated with varying degrees of significance.^7–9, 13, 14^ However, most of them studied only a few variants in underpowered cohorts of less than 100 patients. And none of the described genetic factors could be reproduced in large studies. These targeted approaches may have missed genetic polymorphisms that can identify the patients unlikely to respond to the anti-TNFα biologics that more unbiased approach such as genome-wide association study (GWAS) captures. We conducted the first GWAS to identify the genetic loci associated with PNR to anti-TNFα therapy. The significantly associated locus uncovered by this analysis was replicated in an orthogonal cohort and then tested for any association with a wide range of clinical conditions via a phenome-wide association study (PheWAS) to investigate potential clinical relevance. We also studied the effect of the associated variant on gene expression and TNFα secretion by cell-based assays.

## Methods

### Study participants

All participants provided written informed consent as part of the University of Chicago institutional review board approved protocols. Participants were selected based on (i) an established diagnosis of IBD by a gastroenterologist (CD, Indeterminate Colitis [IC] and Ulcerative Colitis [UC]) by clinical, radiological, and endoscopic examination via chart review (ii) treatment initiation with anti-TNFα biologics (Remacade® (infliximab), Humira® (adalimumab) and CIMZIA® (certolizumab)) (iii) European genetic ancestry. Patients were excluded if (i) they were Non-European ancestry (ii) they were 18 years of age or younger at treatment (iii) Prior therapy with anti-TNF agents (iv) duration of anti-TNF therapy could not be determined through chart review.

### Determination of anti-TNF response phenotype in the discovery and replication cohort

The GWAS discovery and replication cohorts were recruited from the outpatient GI clinic from February 2009 to June 2012. Demographic and health information of the patients were collected from the University of Chicago Hospitals through retrospective review of the electronic heath records. Endoscopic and clinical findings after induction therapy (8 weeks for UC or 12 weeks for CD) were used to determine response to therapy. Patients were categorized as primary non-responders when (i) there was no clinical remission (as noted by the gastroenterologist as marked reduction in diarrhea and abdominal pain, or in the case of patients with fistulae, a decrease in the drainage, size, or number of fistulae for CD, and marked reduction in the amount of diarrhea, hematochezia, and abdominal pain for UC) or (ii) if active inflammation via colonoscopy was present (lack of endoscopic healing) after induction therapy with first exposure to an anti-TNFα biologics. If the dose of the anti-TNFα biologic was increased after induction we allowed an additional 8-12 week to determine response status. Patient had to be discontinued on therapy to be deemed primary non-responders. Responders were patients with favorable clinical and endoscopic response to anti-TNFα induction followed by a continued clinical response after 8-12 weeks of induction therapy in the absence of steroid therapy. Patients that stopped therapy prior to the end of induction due to anaphylactic reactions were excluded from the study. Secondary non-response was defined as loss of response during maintenance after successful induction. If anti-drug antibodies were detected through routine clinical care after loss of response, we categorized these patient as secondary non-responders. However, antibody levels were not available on all subjects. Because of the retrospective nature of our study, secondary non-responders were included as responders.

### Genotyping

DNA of IBD patients receiving anti-TNF therapy was collected through the biobank of the Translational Research Initiative of the Department of Medicine (TRIDOM) of the University of Chicago Hospitals. Patients, who were enrolled after whole-genome genotyping was conducted in the GWAS discovery cohort, served as an independent replication cohort. The discovery cohort of 676 self-reported white patients was genotyped with the Illumina Infinium OmniExpressExome-8 (Illumina, San Diego, CA, USA) at the RIKEN Center for Genomic Medicine (Yokohama, Japan). ^15^ Single nucleotide polymorphisms (SNPs) were excluded based on genotyping rate <95%, minor allele frequency (MAF) <5%^16^ and failed Hardy-Weinberg equilibrium tests p <0.00001. SNPs were also excluded if they were: A/T or C/G SNPs to eliminate flip-strand issues, SNPs on the X and Y chromosomes or mitochondrial SNPs. Samples with a call rate of <95%, missingness > 0.05, discordance between reported gender and chromosomal sex, or identity-by-descent > 0.125 were removed from analysis. Additionally, principal components (PC) 1 and 2 were used to determine genetic ancestry of all individuals with any outliers removed. Genotypes were phased using SHAPEIT and imputed with IMPUTE2 using reference files from the 1,000 Genomes haplotypes -- Phase III integrated variant set release (v3) in NCBI build 37 (hg19) coordinates.^17, 18^ Post imputation quality control involved exclusion of SNPs if the MAF was <0.05, imputation quality <0.8, and failed Hardy-Weinberg equilibrium tests p <0.00001. A total of 589 patients and 6,489,541 variants passed quality control filters and were used for analysis (Supplementary Figure 1). The discovery cohort comprised of 104 primary non-responders and 485 responders to anti-TNF therapy. The replication cohort was genotyped by pyrosequencing and comprised of 32 primary non-responders and 261 responders to anti-TNF therapy.

### Cell Culture, transfection and RNAi

Commercially available lymphoblastoid cell lines (LCLs) from 1000 genome project CEU population (purchased from Coriell Institute) were used to evaluate the effect of the SNP on the *FAM114A2* expression and the protein level. Samples were selected based on their genotype at rs34767465 (GM07000, GM12004, GM12005, GM06985, GM11830, GM11831, GM12760, GM12044, GM12413). LCLs were grown in RPMI-1640 medium containing 10% heat inactivated fetal bovine serum (FBS) supplemented with 10 mM HEPES.

To investigate the functional effect of *FAM114A2* on TNFα secretion, human monocyte THP-1 cells (ATCC, TIB-202) were grown in RPMI-1640 medium containing 10% heat inactivated FBS supplemented with 10 mM HEPES and seeded in 6-well plates at a density of 3.0×10^6^/well before transfection. FAM114A2 siRNAs (SR307384, OriGene) and scrambled control were transfected using Lipofectamine RNAiMAX Reagent (13778030, ThermoFisher) according to the manufacturer’s instruction. Following knockdown, THP-1 monocytes were differentiated into macrophages by incubation with 150 mM phorbol 12-myristate 13-acetate (PMA, Sigma, P8139) for 24 h.^19^

### TNFα ELISA

Human TNFα ELISA Kit (ThermoFisher Scientific, KHC3011) was used to determine the concentration of TNFα in the cell culture supernatant of THP-1 cells-derived macrophages, according to the manufacturer’s protocol.

### Immunoblotting

LCLs and THP-1 cells-derived macrophages were lysed in RIPA lysis buffer (Thermo Scientific, 11965092) supplemented with protease inhibitor cocktail (Roche, 11836170001). Whole cell lysates were centrifuged, and protein concentration was determined using Pierce Coomassie Protein Assay kit (Thermo Scientific, 23200). Equal amounts of proteins were separated by standard SDS/PAGE and transferred to PVDF membrane. The following antibodies were used to quantify protein levels: FAM114A2 (PA5-57441, ThermoFisher), β-tubulin (#86298, CST), GAPDH (MAB374, Millipore), LC3 (2775, Cell Signaling), and p62 (5114, Cell signaling). The protein quantification was performed by Gels Analyze tool of Image J. Representative results obtained from at least three independent experiments were presented.

### Phenome-Wide Association Study (PheWAS)

rs34767465 was tested for its association across a wide range of diseases and traits defined by the phecode system^20^ in eMERGE phase 3 cohort.^21^ Out of 83,717 eMERGE patients, 81,920 patients had both the genotype and phecodes, and were used for this study. Case-control status for each individual was defined for each phecode. Individuals with □≥□3 instances of a phecode were considered as a case, and control status was assigned based on the absence of a given phecode. Samples with ≥1 to <3 phecodes were removed from analysis for any given phecode. A total of 1552 phecodes that occurred in a minimum of 25 cases were tested as previously described (Supplementary Figure 1).^22^

### Statistical Analyses

Differences in baseline characteristics between cases and controls were assessed by χ2 test or Fisher exact test for categorical variables and t-test for continuous parameters. To identify variants associated with anti-TNFα PNR, a case-control GWAS was conducted in the discovery cohort and significant SNPs were tested in the replication cohort. Summary statistics from the discovery and replication cohorts were analyzed in a fixed-effect meta-analysis using METAL (version-2011)^23^, which assumes equal genetic effect between the two cohorts. A one-sided p-value of 5×10^−8^ was considered the Bonferroni corrected significance threshold for the discovery cohort^24^ and meta-analysis. In the replication cohort, a one-sided p-value<0.05 was considered significant as only one SNP was tested in replication. Association analysis was conducted by logistic regression using an additive model. Heterogeneity of the associations across the two cohorts was assessed by Cochran’s Q statistics. Genetic analysis was conducted using SNPTESTv2.5.2 for the discovery cohort and PLINK (version-1.9) for the replication cohort.^25, 26^ The gene region plot for the associated SNP was generated with LocusZoom (version-0.4.8).^27^ For PheWAS analysis, logistic regression was used to test the associations between the selected genetic variant and each phecode after adjusting for sex, decade of birth, eMERGE network recruitment site and the first two principal components. Missing data were handled by the listwise deletion method. Statistical analyses were performed using R (version-3.5.2) software. All authors had access to the study data and reviewed and approved the final manuscript.

## Results

Demographic and clinical characteristics of patient and control groups are shown in Table 1. Age and sex were similar between cases and controls in both the discovery and replication cohorts. Among the clinical covariates collected, use of antibiotics (p<0.0001), methotrexate (p = 0.018) and azathioprine (p = 0.018) were significantly associated with PNR to TNFα therapy in the discovery cohort. Therefore, genome-wide association analyses were adjusted for these covariates. PC1 (p=0.01) was associated with the anti-TNF non-response phenotype and used as a covariate to correct for population substructure.

**Table 1.** Demographic and clinical characteristics of the study groups.

| Variables | Discovery Cohort |  | Replication Cohort |  |
| --- | --- | --- | --- | --- |
|  | Cases<br>(n = 104) | Controls<br>(n = 485) | Cases<br>(n = 32) | Controls<br>(n = 261) |
| Age, years<br>(mean ±SD) | 36.62±14.39 | 35.88±14.23 | 36.53±16.88 | 30.29±15.14 |
| Sex (%) Female | 50 (48.1) | 232 (47.8) | 13 (40.6) | 137 (52.5) |
| Male | 54 (51.9) | 253 (52.2) | 19 (59.4) | 124 (47.5) |
| Smoking n (%) | 40 (38.5) | 187 (38.1) | 15 (46.9) | 83 (31.8) |
| GI dysplasia/ Cancer n (%) | 5 (4.8) | 34 (7) | 3 (9.4) | 14 (5.4) |
| Calcineurin inhibitor n (%) | 2 (1.9) | 6 (1.2) | - | 3 (1.1) |
| *Antibiotics n (%) | 26 (25) | 225 (46.4) | 13 (40.6) | 115 (45.1) |
| *Methotrexate n (%) | 9 (8.7) | 88 (18.1) | 5 (16.1) | 65 (25) |
| *Azathioprine n (%) | 46 (44.2) | 276 (56.9) | 11 (34.4) | 128 (49.8) |
| Aminosalicylates n (%) | 33 (32) | 140 (29.2) | 11 (35.5) | 60 (23.6) |
| **Anti-TNFα therapy** |  |  |  |  |
| Infliximab n (%) | 66 (64.1) | 328 (67.9) | 14 (43.8) | 154 (59) |
| Adalimumab n (%) | 29 (28.2) | 146 (30.2) | 17 (53.1) | 98 (37.5) |
| Certolizumab n (%) | 9 (8.6) | 11 (2.3) | 1 (3.1) | 9 (3.4) |
| **IBD type** |  |  |  |  |
| Crohn's Disease n (%) | 73 (70.2) | 387 (80) | 28 (87.5) | 255 (98) |
| Ulcerative Colitis n (%) | 28 (26.9) | 85 (17.5) | - | - |
| Indeterminate Colitis n (%) | 3 (2.8) | 13 (2.6) | 4 (12.5) | 6 (2.3) |

\* p <0.05, statistically significant in the discovery cohort

### GWAS findings

In the discovery cohort, rs34767465 on chromosome 5, along with 9 SNPs in high linkage disequilibrium with this lead SNP, was significantly associated with PNR to anti-TNFα therapy (OR: 2.07, 95% CI: 1.46-2.94, p = 2.43 × 10^−7^) (Table 2, Figure 1A). While 7 of the 9 SNPs were imputed, two of them (rs13173354 and rs9324761) were present on the genotyping platform. rs201833877 on chromosome 11 was also associated with PNR (OR: 3.17, 95% CI: 1.87-5.36, p = 7.18 ×10^−7^), however the variant is an insertion/deletion within a long repeat sequence and could not be replicated by pyrosequencing. Association of rs34767465 was confirmed in the replication cohort (OR: 1.8, 95% CI: 1.04-3.16, p = 0.03). Near genome-wide significance of this SNP was achieved when the cohorts were combined via meta-analysis (meta-analysis p = 5.2×10^−8^), with no significant heterogeneity between cohorts detected (Cochran Q statistic P□=□0.21). rs34767465 is an intergenic variant located between *FAM114A2* and *GALNT10* (Figure 1B). Data from the GTEx consortium (v8) reports rs34767465 as a *cis* expressed quantitative locus (cis-eQTL) for *FAM114A2* in skeletal muscle (Figure 1C) while data from NESDA NTR conditional eQTL catalog ^28–30^ shows that rs34767465 is also a *cis*-eQTL for *FAM114A2* in whole blood.

**Table 2:**
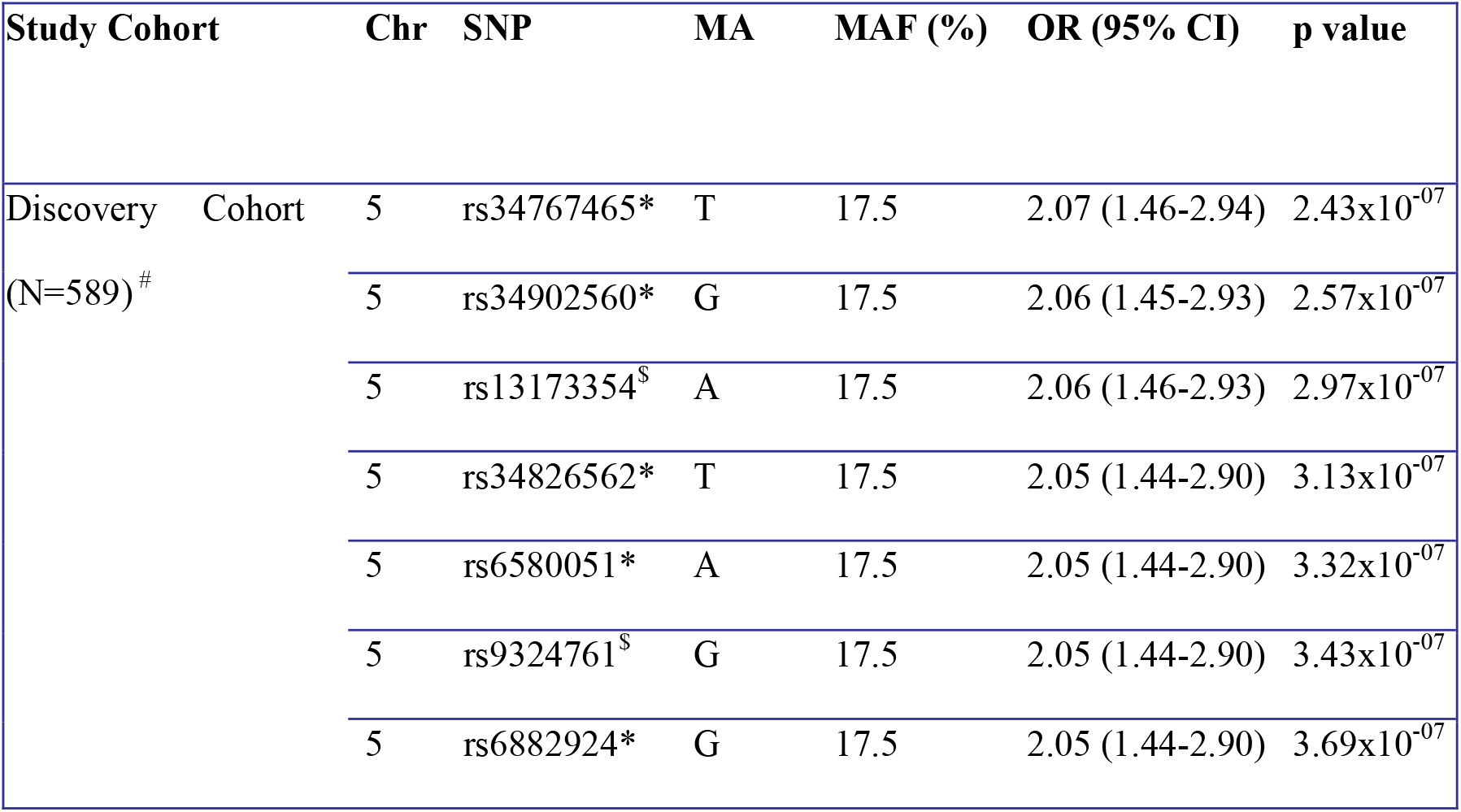
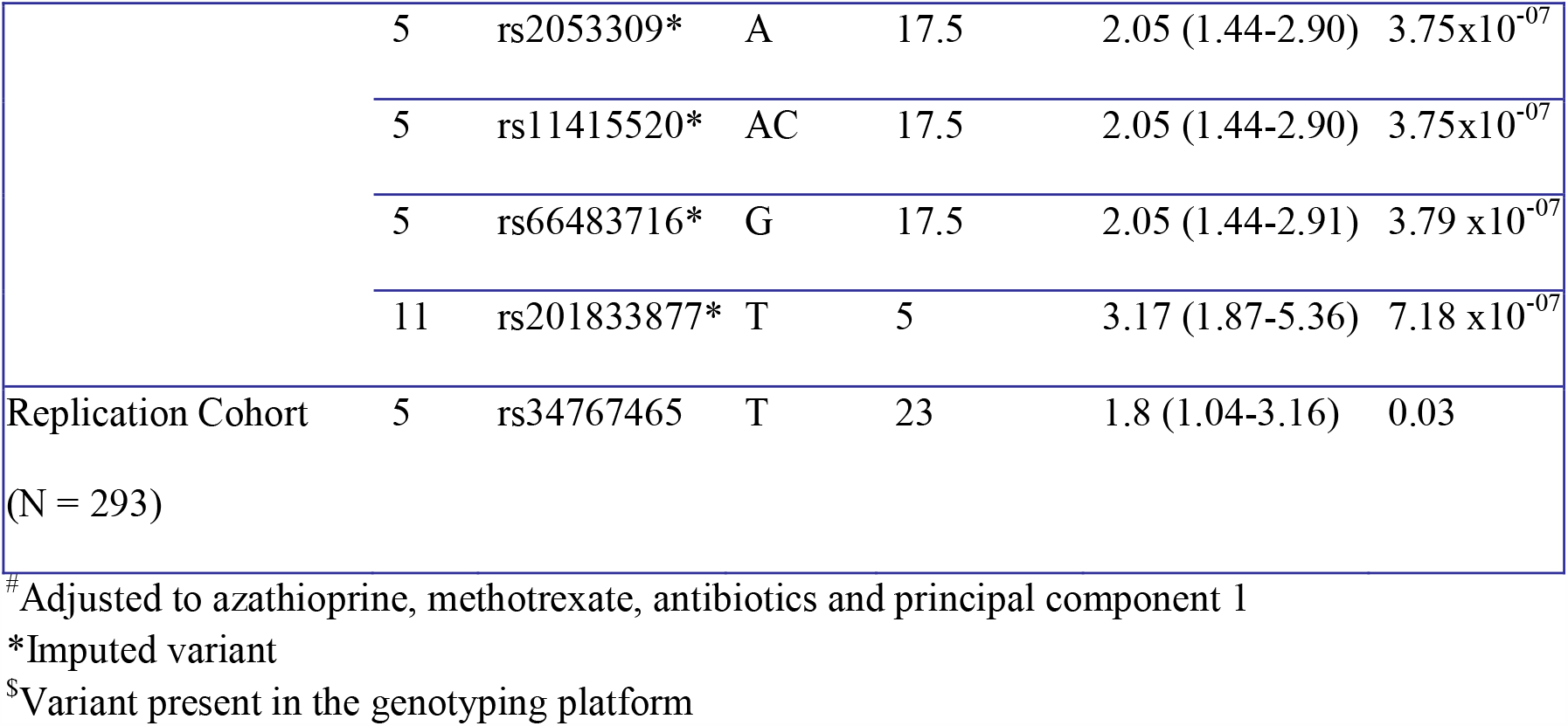
GWAS loci for non-response to anti-TNF biologics.

**Figure 1:**
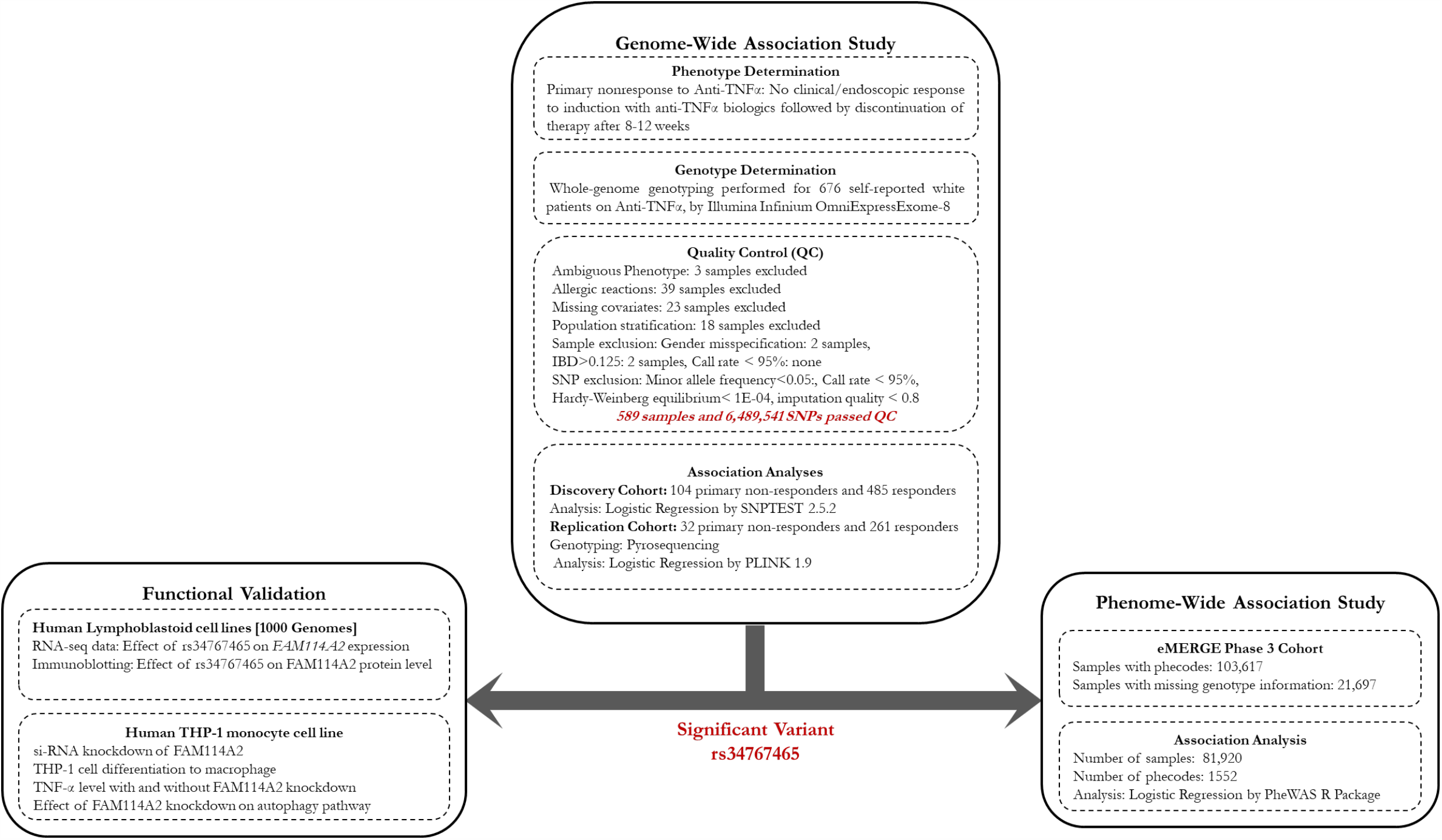
Study outline and analyses workflow. The figure outlines the genome-wide association study that identified and replicated a genetic locus rs34767465 associated with primary non-response to anti-TNFα therapy. rs34767465 was then tested for any association with a wide range of clinical conditions via a phenome-wide association study to investigate potential clinical relevance. rs34767465 was also studied for its effect on gene expression and TNFα secretion by cell-based assays.

We sought to determine if the association of rs34767465 with PNR was influenced by the IBD sub-types. GWAS restricted to only CD patients in the GWAS cohort (Case: 73, Controls: 387) showed an association of 1.8-fold risk of PNR (OR: 1.8, CI: 1.18-2.76, p = 1.1 × 10^−3^), while in only UC patients in the GWAS cohort (Case: 28, Controls: 85) the association was 2.89-fold (OR: 2.89, CI: 1.45-5.78, p = 4.1 × 10^−4^). These findings show that rs34767465 may have differential effects by disease sub-types. However, the p-value was below the genome-wide statistical significance threshold due to the small number of samples in either subtype.

We also looked into the previously reported SNPs associated with PNR in the discovery cohort and observed no association (Supplementary Table 1).

### Effect of rs34767465 on *FAM114A2* gene expression

LCLs from 1000 Genomes Project were used to evaluate the effect of rs34767465 on the gene expression of *FAM114A2*. RNA sequencing (RNA-seq) from 89 HapMap CEU LCLs (62 of wild type, 24 of heterozygous, 3 of homozygous) showed that the mRNA level of *FAM114A2* was decreased with 14% decline in heterozygous gene expression and 14% decline in homozygous gene expression as compared to wild type (Figure 2A). However, we only saw a statistically significant decrease in heterozygous LCLs, not in homozygous LCLs due to small sample size (n=3). Immunoblotting showed that the FAM114A2 protein level declined with rs34767465 genotypes with 40% decrease in heterozygous (Het) protein expression and 68% decrease in homozygous risk allele (Hom) protein expression as compared to wild type (WT); consistent with the gene expression data (Figure 2B and 2C).

**Figure 2a:**
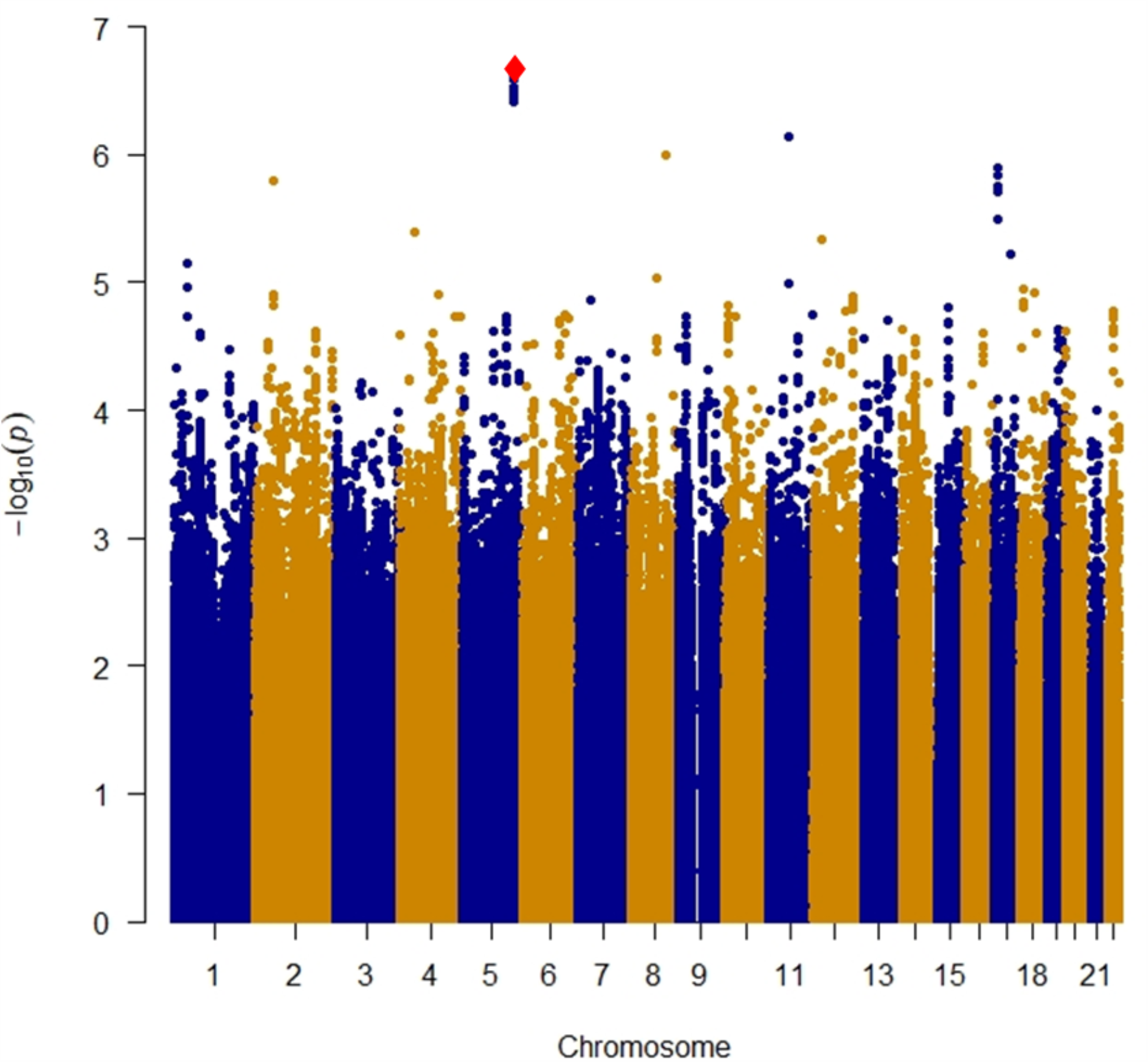
Manhattan plot of near-significant loci associated with primary non-response to anti-TNFα biologics in the discovery cohort. Single-nucleotide polymorphisms (SNPs) are plotted on the x-axis according to their positions on each chromosome against association with primary non-response to anti-TNFα biologics on the y-axis (−log10*P* value). Near-significant associations were observed in chromosome 5 [Genome-wide significant threshold: *P* = 5.0 × 10^−8^]. The red diamond identifies our most significant SNP association (rs34767465).

**Figure 2b:**
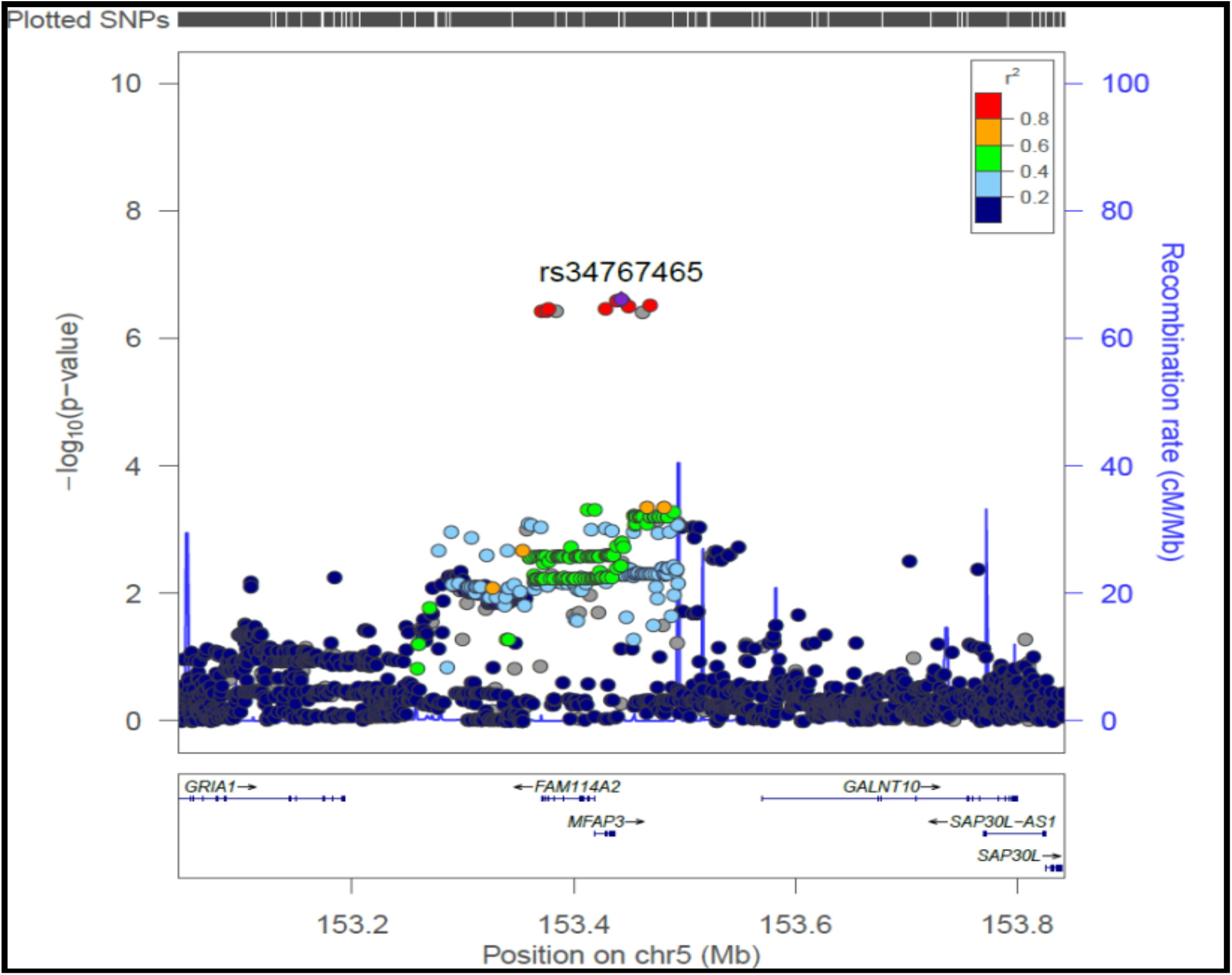
Locus-specific plot of rs34767465 on chromosome 5. The x-axis represents the genomic position in chromosome 5 and the left y-axis represents the – log10*P* of association with primary non-response to anti TNF∝ biologics in the discovery cohort. The colors of the circles denote linkage disequilibrium (r^2^) between rs34767465 (purple diamond) and nearby SNPs (based on pairwise *r*^*2*^ values from the 1000 Genomes Project European population). The right y-axis show the estimated recombination rate (obtained from HapMap). Genes at this locus are indicated in the lower panel of the plot. Chromosomal positions are based on hg19 genome build.

**Figure 2c:**
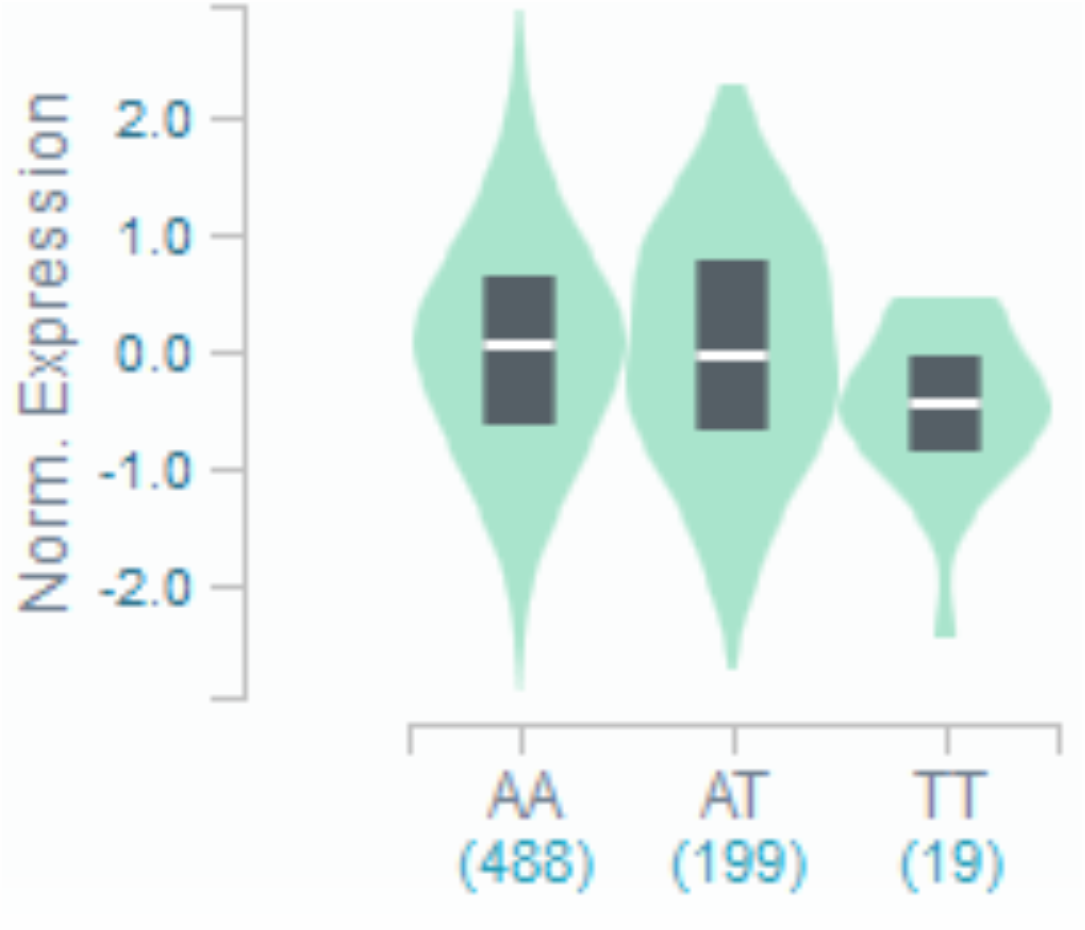
Violin plot of *FAM114A2* expression for *cis* eQTL rs34767465 in Genotype-Tissue Expression (GTEx, release v7) The allelic effect of rs34767465 on normalized *FAM114A2* gene expression levels are shown by boxplots within violin plots. [Normalized effect size = −0.15, p = 0.0000027]. A and T alleles indicate the major and minor allele types, respectively with the number of subjects shown under each genotype. The plots indicate the density distribution of the samples in each genotype. The white line in the box plot (black) shows the median value of the gene expression at each genotype.

### Effect of *FAM114A2* knockdown on TNFα secretion

To investigate the effect of *FAM114A2* on TNFα secretion, *FAM114A2* siRNAs were transfected into the THP-1 cells to knock down *FAM114A2* gene expression and then THP-1 cells were differentiated into macrophages using PMA. A significant increase of 24% and 41% in relative TNFα secretion was observed with *FAM114A2* knockdowns (Figure 3A). To further investigate the mechanism of *FAM114A2* in the secretion of TNFα, we examined the involvement of *FAM114A2* in autophagy, which plays an important role in the inflammatory pathology of IBD. THP-1 cells were transfected with scrambled control and *FAM114A2* siRNAs for 48h. Immunoblotting showed that knockdown of *FAM114A2* decreased the expression of both LC3 subtypes (89% of LC3-I, 24% of LC3-II, and 86% of the ratio of LC3-I/ LC3-II) and 100% increase in the relative expression of P62 protein expression, suggesting depletion of *FAM114A2* impaired autophagy related pathways, thereby promoting TNFα secretion. (Figure 3B and 3C).

**Figure 3:**
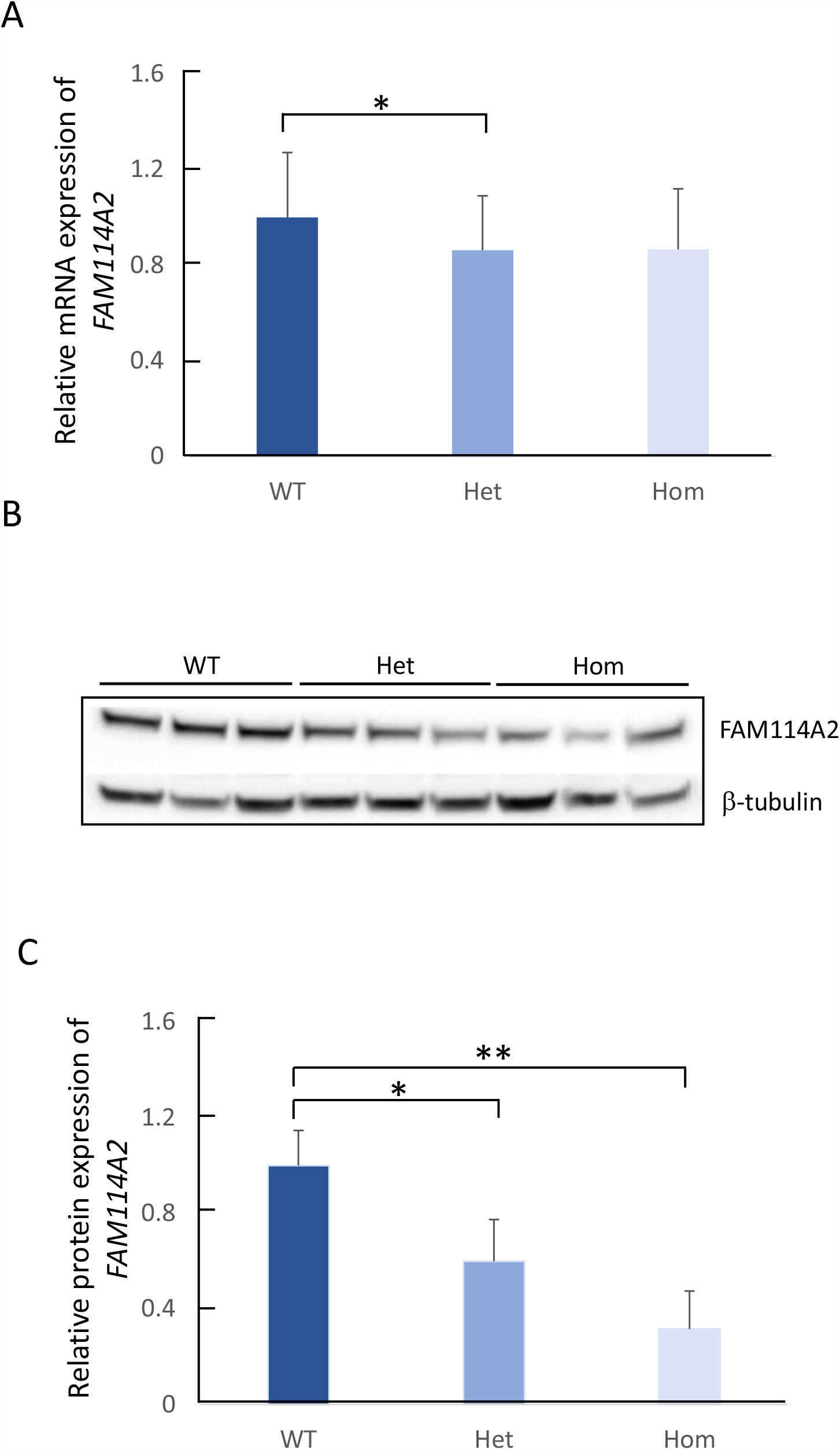
The effect of rs34767465 on *FAM114A2* gene and protein expression in LCLs. a) Relative gene expression of *FAM114A2* by rs34767465 genotypes in LCLs (number of subjects per genotype group - WT = 62, Het = 24, Hom = 3). b) Immunoblot gel image showing decreased FAM114A2 protein expression by rs34767465 genotypes. Three different LCL lines with known genotypes were used. c) Quantitative analysis of immunoblotting results using Image J. Protein values are normalized by β-tubulin expression and are shown relative to WT expression. Wild type-WT, Heterozygous-Het, Homozygous-Hom, LCL-Lymphoblastoid Cell lines. * p<0.05 ** p<0.01.

**Figure 4:**
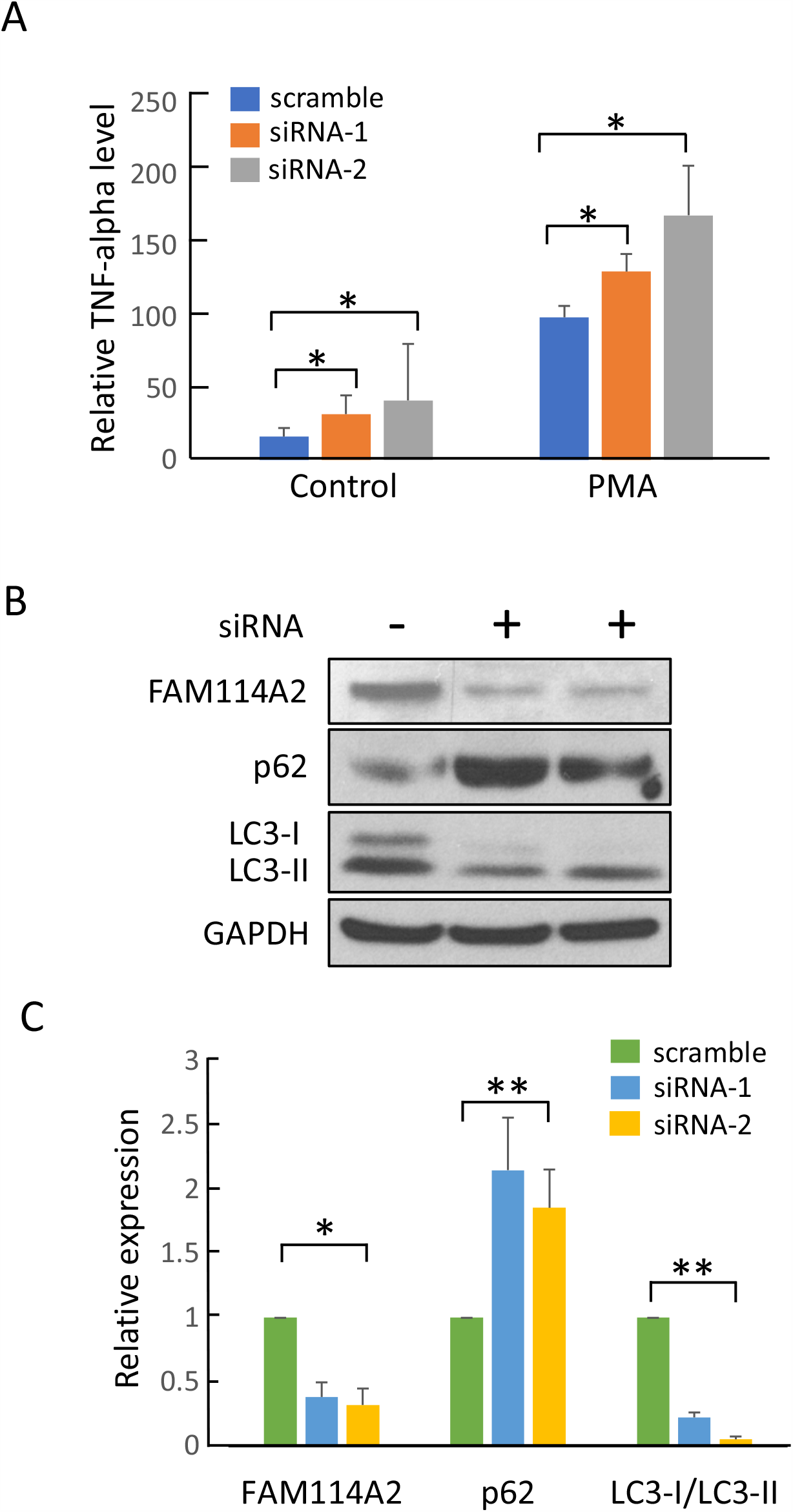
Functional analysis of *FAM114A2* in THP-1 cells. a) *FAM114A2* siRNAs and scramble were transfected into THP-1 cells and untreated control, following which THP-1 cells were treated with PMA (150mM) for 24 hours to transform the cells to macrophages. Cell culture supernatant was collected for TNFα ELISA analysis. All TNFα level values are shown as relative to PMA treated scramble control. All experiments were conducted in triplicate. b) Immunoblot analysis showing *FAM114A2* gene knockdown increased the autophagy related protein, P62, and decreased the LC3 protein level - Representative Immunoblotting image shown. c) Quantitative analysis of Immunoblotting results using Image J. All protein expression values were normalized to GAPDH and are shown relative to scramble control in each group. All experiments were conducted in triplicate. phorbol 12-myristate 13-acetate – PMA, * p<0.05 ** p<0.01.

**Figure 5:**
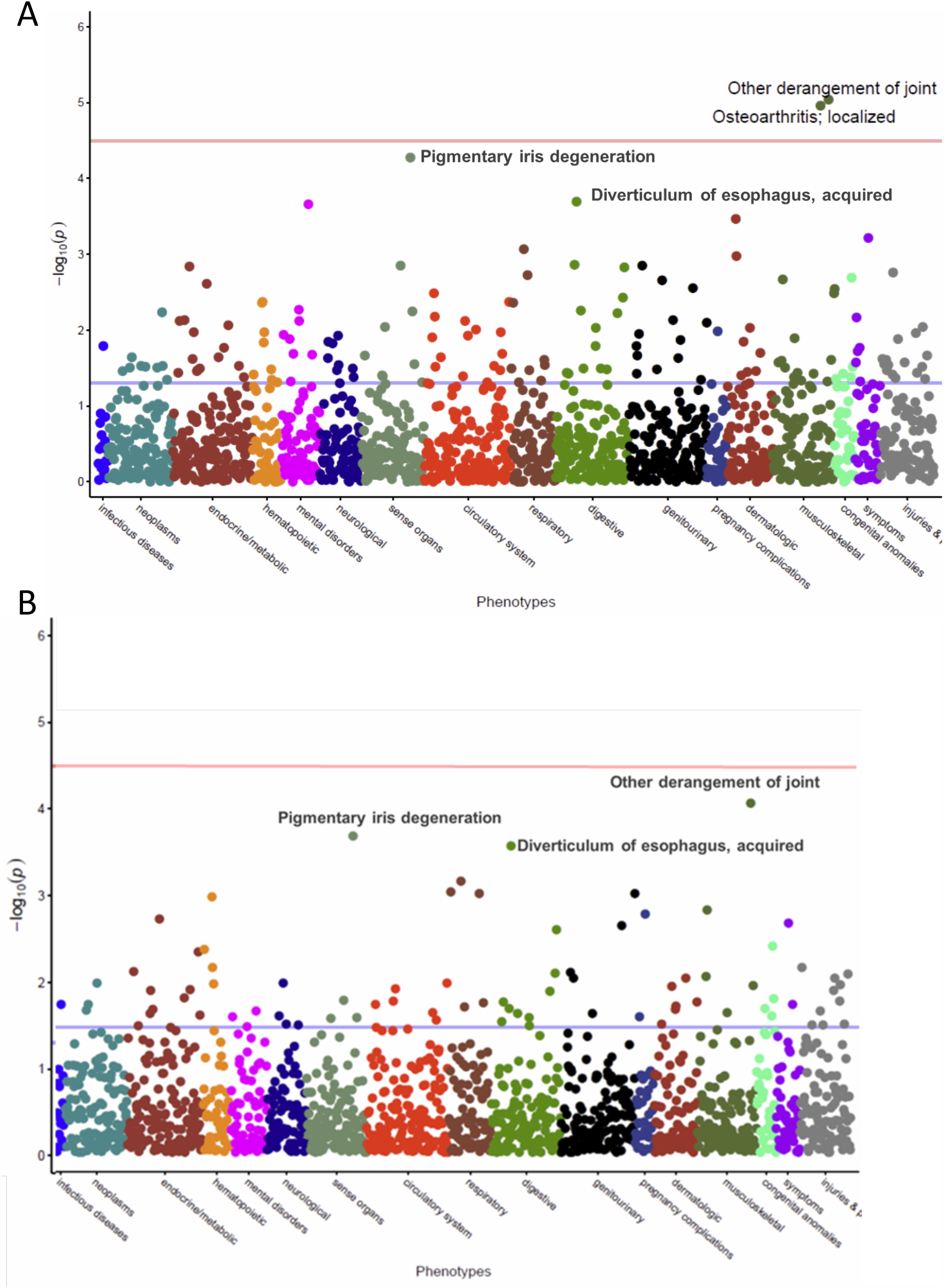
PheWAS Manhattan plot for phenotypes associcated to rs34767465. Plots represent 1,552 phenotypes tested for association with rs34767465. Phenotypes grouped along the x axis by categorization within the PheWAS code hierarchy. The Y axis reflects the P value for each phenotype. The blue horizontal line represents nominal p-value threshold of 0.05, the red horizontal line represents Bonferroni corrected p-value threshold of 3.2 × 10^−5^, respectively. a) PheWAS analysis adjusted to decade of birth, gender, PC1, PC2 b) PheWAS analysis adjusted to decade of birth, gender, PC1, PC2 and eMERGE recruitment site

### PheWAS findings

The mean age of the PheWAS cohort was 49.2 ± 24.9 years and 44165 (53.9%) were females. rs34767465 was strongly associated with derangement of joints (OR: 1.3, p =9 × 10^−6^) and osteoarthritis (OR: 1.14, p = 1.1 × 10^−5^), when adjusted for decade of birth, gender and principal component (PC) 1 and 2 (Supplementary Figure 2A, Table 3). The other phenotypes that were close to Bonferroni-corrected significance were pigmentary iris degeneration (OR: 1.73, p= 5.1 × 10^−4^) and acquired diverticulum of the esophagus (OR: 1.88, p=6.3 × 10^−4^). None of the phenotypes crossed Bonferroni-corrected significance level (p = 3.2 × 10^−5^) after adjusting for eMERGE recruitment site along with decade of birth, gender, PC1 and PC2 (Supplementary Figure 2B).

**Table 3:**
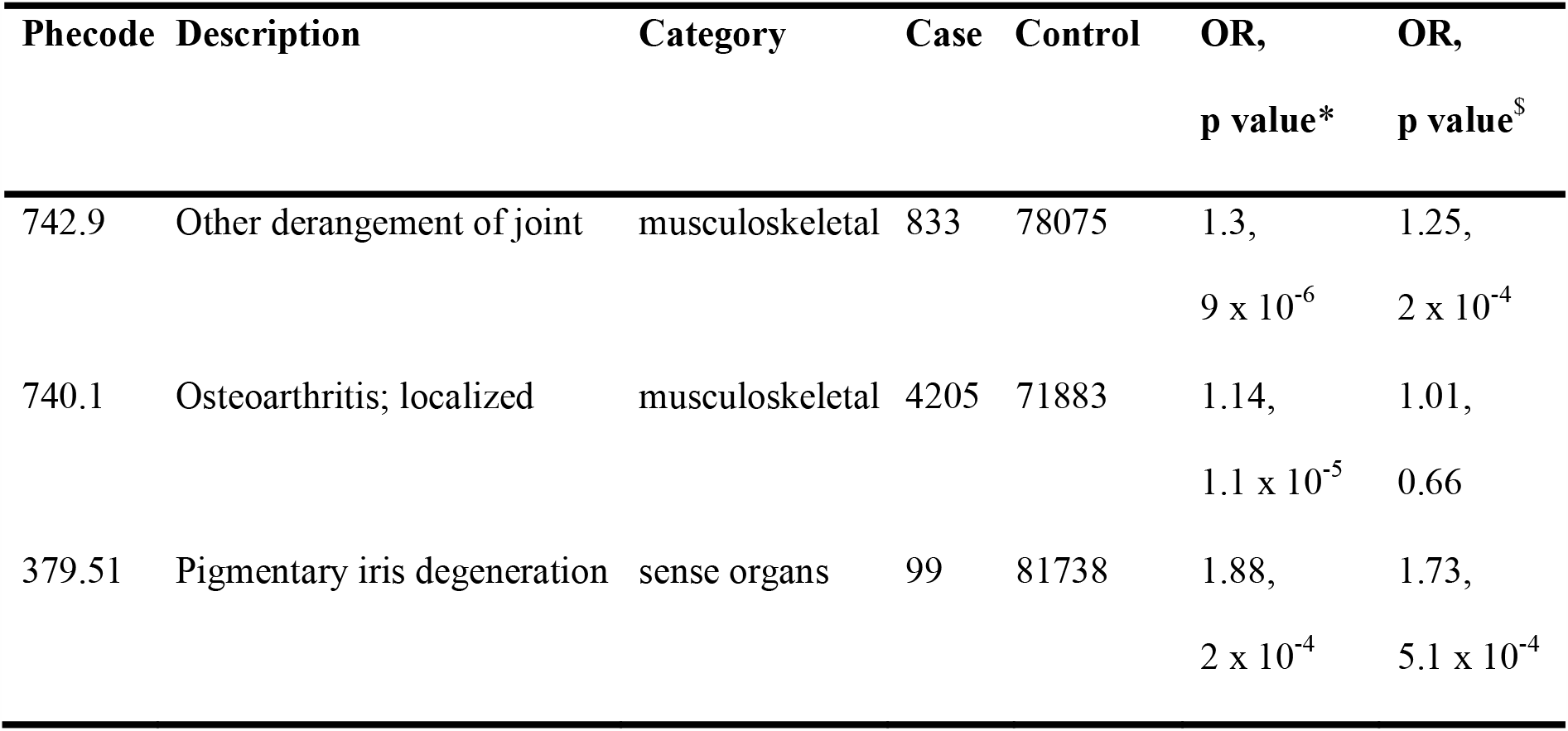
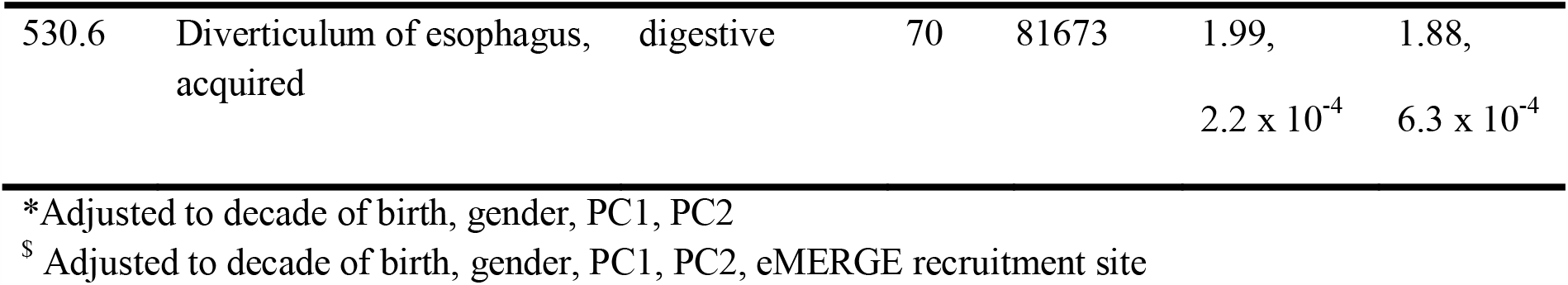
PheWAS analyses for significant loci rs34767465.

## Discussion

This is the first GWAS to investigate the genetic basis of PNR to anti-TNFα treatment in patients with IBD. We identified a novel association with common variant rs34767465, found to impart 2-fold increased risk of PNR to anti-TNFα therapy. Based on GTEx data and the NESDA NTR conditional eQTL catalog, this SNP affects the expression of *FAM114A2* in skeletal muscle tissue and whole blood. We confirmed the effect of the rs34767465T allele on reduced expression of *FAM114A2*, at both the mRNA and protein level, in LCLs obtained from 1000 genomes CEU population. Although the function of *FAM114A2* is not well known, a phosphoproteomic study showed that Leucine-rich repeat kinase 2 (LRRK2) kinase inhibition led to ≥50% alteration in the phosphorylation of FAM114A2 protein.^31^ *LRRK2* is a major susceptibility gene for Crohn’s disease (CD) and deficiency of the gene in mice confers enhanced susceptibility to experimental colitis.^32^ We did not observe significant difference in the *LRRK2* mRNA or protein levels by knockdown of *FAM114A2* and vice versa, in LCLs (data not shown). However, due to lack of availability of phosphoprotein antibody for FAM114A2, the effect of LRRK2-mediated phosphorylation of FAM114A2 could not be studied. Our study showed that *FAM114A2* knockdown in THP-1 cells-derived macrophages led to an impaired autophagy pathway, thereby increasing TNFα secretion. Dysfunctional autophagy plays an important role in IBD pathogenesis by altering processes like defense against infections, carcinogenesis, antimicrobial peptide secretion by Paneth cells, goblet cell function, pro-inflammatory cytokine production by macrophages, antigen presentation by dendritic cells and the endoplasmic reticulum stress response in enterocytes.^33^ Our findings revealed that FAM114A2 may regulate TNFα levels by inducing the autophagy pathway. However, IBD patients carrying the rs34767465 T allele have reduced *FAM114A2* expression leading to impaired autophagy and increased TNFα secretion by macrophages. These patients therefore may not respond to the standard dose of anti-TNFα and tailored dosing might be helpful for an optimal response. Suboptimal drug concentrations are associated with clinical lack or loss of response, leading to treatment failure and drug discontinuation. A study by Kennedy *et*.*al*., investigated the predictors of anti-TNF treatment failure, and reported that the only factor independently associated with PNR was low drug concentration. The study demonstrated that adequate drug concentrations at week 14 were associated with clinical remission, decreased risk of developing anti-drug antibodies and better long-term outcomes.^34^ Therefore, ensuring adequate drug concentrations is of utmost importance during both the initiation phase to prevent PNR as well as the maintenance phase to avoid secondary loss of response. Given that drug levels are not routinely taken in IBD patients we are not able to assess if non-response could be due to lower concentration of medication.

We also observed a locus on chromosome 11, rs201833877 associated with PNR in the discovery cohort. Based on GTEx data, this variant is a multi-tissue splice quantitative trait loci for *MS4A7*, a gene that is expressed in microglia and brain border macrophages and is associated with immune function.^35, 36^ However, the variant is an insertion/deletion within a long repeat sequence and could not be reliably genotyped in the replication cohort.

Previous studies investigated the genetic basis of PNR to anti-TNFα therapy by targeting genes related to immunological pathways. We observed no association of the reported variants in our discovery cohort. The lack of association could be due to the differences in the definition of PNR that often varies between studies. While some studies defined response based on the CRP levels or calprotectin levels, others looked at the endoscopic response or clinical response such as UC Disease Activity Index and CD Activity Index. However, these measures are not always assessed in routine clinical care of IBD and therefore were not available for most of our cohort.

PheWAS analysis of rs34767465 identified pleotropic associations with comorbidities like derangement of joints, osteoarthritis, pigmentary iris degeneration and diverticulum of the esophagus. Although these phenotypes did not reach the Bonferroni corrected significance threshold after adjusting for all the covariates, these conditions are known to co-occur in patients with IBD. A connection of IBD and diverticular disease has long been recognized. Crohn’s disease and diverticulitis share clinical and radiologic features and studies suggest that even the characteristic pathology of Crohn’s disease can be a secondary reaction to diverticulitis.^37^ Sultan *et al*., examined the relationship of colonic diverticulosis with IBD and observed an increased frequency of sigmoid and rectal inflammation, extraintestinal manifestations (EIMs), and an older age of IBD onset in cases with diverticulosis. The study speculated that diverticula may lead to changes in the colonic microflora that can in turn promote IBD.^38^ However, the phenotype diverticulum of esophagus did not achieve the significance threshold in our PheWAS analysis, which could be due to the low number of cases for this phenotype. A variety of chronic comorbidities have been associated with IBD, as the disease might share pathogenic pathways with other conditions. The most common EIMs in IBD are musculoskeletal disorders, often associated with colonic involvement, and present as either articular (arthritis) or periarticular inflammation including enthesitis, myositis, or soft tissue rheumatism (fibromyalgia).^39^ A study showed that peripheral arthritis in IBD patients is influenced by major histocompatibility complex haplotypes HLA-B27, HLA-B35, HLA-DR, and HLA-B44, and therefore could be genetically driven.^40^ Ophthalmologic problems can occur as an EIM of the disease or may be drug-related.^41^ A study by Santeford *et al*, demonstrated that autophagy is essential for maintaining ocular immune privilege, meaning the eye’s ability to limit local immune and inflammatory responses to preserve vision, and deletion of multiple autophagy genes in macrophages leads to an inflammation-mediated eye disease.^42^ Based on these findings it can be speculated that rs34767465 not only influences IBD pathogenesis leading to non-response to anti-TNFα therapy but can also explain the associated comorbidities that are often observed in IBD patients. Therefore, the SNP may help guide physicians’ drug choice and therapeutic drug monitoring to prevent PNR to anti-TNFα therapy and IBD associated comorbidities.

## Limitations

This study has several limitations. First, the small sample size prevented genome-wide significance. However, association of the SNP with PNR was confirmed in an independent replication cohort. Also, the SNP achieved near genome-wide significance threshold on meta-analysis of the combined discovery and replication cohorts. Second, although rs201833877 on chromosome 11 was also associated with PNR to anti-TNFα therapy, the variant is an insertion/deletion of a long repeat sequence and could not be replicated. Third, we could not establish the link between LRRK2 and FAM114A2 due to lack of availability of a phosphoprotein antibody. Fourth, we did not cross the Bonferroni-corrected significance threshold for interesting phenotypes in the PheWAS analyses, which could be due to low number of cases, seen in the diverticulum of esophagus phenotype. Lastly, there was a lack of information on plasma biomarkers like anti-TNFα concentration, CRP levels or anti-drug antibody level for our patient cohorts. Although studies suggest the utility of these parameters in determining PNR,^43, 44^ these are not part of routine clinical care in IBD therapy and therefore were not available for most of the patients. Hence, we are not able to account for these variables in our analysis.

## Conclusion

This is the first GWAS to identify a significant variant associated with PNR to anti-TNFα biologic therapy and may explain the comorbidities associated with IBD. The findings may help guide physicians’ drug choice and therapeutic drug monitoring to prevent PNR to anti-TNFα therapy and IBD-associated comorbidities.

## Data Availability

Data are available on reasonable request. All data relevant to the study are included in the article.

## Acknowledgement

The authors thank PGRN-CGM International Collaborative Studies for providing genotyping service.

---

**Figure 1**

**A): Manhattan plot of near-significant loci associated with primary non-response to anti-TNF**α **biologics in the discovery cohort**.

Single-nucleotide polymorphisms (SNPs) are plotted on the x-axis according to their positions on each chromosome against association with primary non-response to anti-TNFα biologics on the y-axis (−log_10_*P* value). Near-significant associations were observed in chromosome 5 [Genome-wide significant threshold: *P*□=□5.0□x□10^−8^]. The diamond identifies our most significant SNP association (rs34767465).

**B): Locus-specific plot of rs34767465 on chromosome 5**

The x-axis represents the genomic position in chromosome 5 and the left y-axis represents the – log_10_*P* of association with primary non-response to anti TNF∝ biologics in the discovery cohort. The colors of the circles denote linkage disequilibrium (r^2^) between rs34767465 and nearby SNPs (based on pairwise *r*^*2*^ values from the 1000 Genomes Project European population). The right y-axis show the estimated recombination rate (obtained from HapMap). Genes at this locus are indicated in the lower panel of the plot. Chromosomal positions are based on hg19 genome build.

**C): Violin plot of *FAM114A2* expression for *cis* eQTL rs34767465 in Genotype-Tissue Expression (GTEx, release v7)**

The allelic effect of rs34767465 on normalized *FAM114A2* gene expression levels are shown by boxplots within violin plots. [Normalized effect size = −0.15, p = 0.0000027]. A and T alleles indicate the major and minor allele types, respectively with the number of subjects shown under each genotype. The plots indicate the density distribution of the samples in each genotype. The white line in the box plot (black) shows the median value of the gene expression at each genotype.

**Figure 2: The effect of rs34767465 on *FAM114A2* gene and protein expression in LCLs**

**A)** Relative gene expression of *FAM114A2* by rs34767465 genotypes in LCLs (number of subjects per genotype group - WT = 62, Het = 24, Hom = 3).

**B)** Immunoblot gel image showing decreased FAM114A2 protein expression by rs34767465 genotypes. Three different LCL lines with known genotypes were used.

**C)** Quantitative analysis of immunoblotting results using Image J. Protein values are normalized by β-tubulin expression and are shown relative to WT expression.

Wild type-WT, Heterozygous-Het, Homozygous-Hom, LCL-Lymphoblastoid Cell lines. b* p<0.05 ** p<0.01.

**Figure 3: Functional analysis of *FAM114A2* in THP-1 cells**.

**A)** *FAM114A2* siRNAs and scramble were transfected into THP-1 cells and untreated control, following which THP-1 cells were treated with PMA (150mM) for 24 hours to transform the cells to macrophages. Cell culture supernatant was collected for TNFα ELISA analysis. All TNFα level values are shown as relative to PMA treated scramble control. All experiments were conducted in triplicate.

**B)** Immunoblot analysis showing *FAM114A2* gene knockdown increased the autophagy related protein, P62, and decreased the LC3 protein level - Representative Immunoblotting image shown.

**C)** Quantitative analysis of Immunoblotting results using Image J. All protein expression values were normalized to GAPDH and are shown relative to scramble control in each group. All experiments were conducted in triplicate.

phorbol 12-myristate 13-acetate – PMA, * p<0.05 ** p<0.01.

